# Analysis of baroreflex activation therapy in patients with heart failure with reduced ejection fraction on current era guideline-directed medical therapy

**DOI:** 10.64898/2026.01.30.26345253

**Authors:** Vu Pham, Arnold Gan, Pratik Doshi, David Valdivia, Melissa Lee Wilson, Michael W. Fong

**Affiliations:** Department of Medicine, Keck School of Medicine of the University of Southern California, Los Angeles, CA; Division of Cardiovascular Medicine, Keck School of Medicine of the University of Southern California, Los Angeles, CA; SC-CTSI Biostatistics, Epidemiology, and Research Design, Keck School of Medicine of the University of Southern California, Los Angeles, CA; Department of Population and Public Health Sciences, Keck School of Medicine of the University of Southern California, Los Angeles CA

## Abstract

**Background:** Guideline-directed medical therapy (GDMT) has been shown to improve mortality and/or symptoms in heart failure with reduced ejection fraction (HFrEF). Medical devices also play an important role in improved quality of life and overall symptom relief for HFrEF patients. Baroreflex Activation Therapy (BAT) increases parasympathetic nervous system activity by stimulating the carotid baroreceptors, thereby reducing symptoms. Herein, we analyzed the effects of BAT on hospitalization, atrial arrhythmia (AA), and ventricular arrhythmia (VA) rates.

**Methods:** A retrospective cohort study was conducted consisting of HFrEF patients treated with BAT at Keck Hospital of USC between 11/2014 and 11/2022. We compared median pre-BAT hospitalization, AA, and VA rates to post-BAT rates at both 6- and 12-months using Wilcoxon Signed Rank tests.

**Results:** Among 31 patients on BAT, 38.7% met criteria for receiving all four GDMT classes for at least 12 months prior to BAT. Among these, 91.7% had an implantable cardioverter defibrillator (ICD) implanted for ≥12 months pre- and post-BAT. Average pre- vs. post-BAT all-cause hospitalization rates were significantly different only at 12 months [1.3 ± 1.4 vs 0.3 ± 0.9, respectively (p=0.05)]. Borderline significant pre-post comparisons were noted including decreased VA rate at both 6 and 12 months and increased AA rate at 12-months (p=0.06 for all).

**Conclusion:** In HFrEF patients on full GDMT, BAT was associated with a significant reduction in hospitalization rates at 12 months. There were no significant changes in AA or VA rates.

## Introduction

With the advancement of guideline directed therapy (GDMT) and medical device therapies for treatment of heart failure (HF), the prevalence of HF is projected to increase by 46% from 2012 to 2030, affecting >8 million people ≥18 years of age due to the aging population.^1^ The lifetime risk of HF remains high, with variation across racial and ethnic groups ranging from 20% to 46% after 45 years of age.^2^ Along with GDMT, medical devices continue to play an important role in improved quality of life and overall symptom relief and will grow in importance as the prevalence of HF continues to rise.

The Baroreflex Activation Therapy for Heart Failure (BeAT-HF) trial focused on Baroreflex Activation Therapy (BAT) in patients with HF and evaluated symptomatic relief showing significant improvement in quality of life, exercise capacity, and NT-proBNP.^3^ A recent analysis of patients with implantable BAT concluded that BAT was associated with significant reductions in all-cause, cardiovascular, and HF related hospitalizations, as well as shorter hospital stays, indicating improved clinical stability in patients with HF.^4^ At the time of the BeAT-HF trial, GDMT consisted of three medication classes which had been shown to improve mortality and/or symptoms specifically in heart failure with reduced ejection fraction or mildly reduced ejection fraction (HFrEF/HFmrEF).^3^ This regimen included beta-blockers, angiotensin converting enzyme inhibitors (ACEi)/angiotensin receptor blockers (ARBs) and mineralocorticoid receptor antagonists (MRA) with each medication titrated as maximally tolerated. However, the most recent GDMT additions of angiotensin receptor-neprilysin inhibitors (ARNI) and sodium-glucose cotransporter 2 (SGLT-2) inhibitors were not fully available at the time of the study.

While the BeAT-HF trial has shown improvement in several metrics, to our knowledge, the impact of BAT has never been studied in conjunction with the current four pillars of GDMT.

Patients with HFrEF have increased risk of severe ventricular arrhythmia (VA), ultimately leading to increased mortality.^5^ VA encompasses a wide range of abnormal cardiac rhythms including monomorphic and polymorphic ventricular tachycardia (VT), ventricular fibrillation (VF), and premature ventricular contractions (PVC).^6^ Additionally, patients with HFrEF have increased risk of atrial arrhythmias (AA), which includes atrial fibrillation (AF), atrial flutter (AFL), and atrial tachycardia (AT).^7^ The sympathetic and parasympathetic nervous system play an important role in the occurrence of VT and VF.^8^ BAT employs an implantable device to reduce sympathetic outflow and increases parasympathetic activity by stimulating the carotid baroreceptors thereby reducing afterload and HF symptoms.^3,9^ However, to our knowledge, the effects of BAT on AA and VA have never been studied.

In this retrospective study, we aimed to investigate the impact of BAT while paired with current-era GDMT primarily to see if there were significant reductions in hospitalizations pre and post-BAT at 6 and 12 months. We secondarily investigated AA and VA rates at the same time points.

## Material and Methods

### Database and cohort

Study approval was obtained through the University of Southern California (USC) Institutional Review Board (ID: HS-23-00546) prior to data collection. Patients with HFrEF on BAT were identified at Keck Hospital of USC and data was obtained through the Cerner electronic medical record (EMR) system. A retrospective cohort study was conducted on patients with HFrEF on maximally tolerated GDMT and BAT. The primary outcome in the study was the difference in hospitalization rates and secondary outcomes were differences in AA and VA rates. Patients on all four pillars of GDMT at least 12 months pre- and post-BAT therapy were included using Day 0 as the date of implant. To measure the secondary outcomes, patients were further categorized into the presence or absence of an implantable cardioverter-defibrillator (ICD) at least 12 months pre- and post-BAT therapy. Patients without an ICD were excluded from the study (Figure 1).

**Figure 1.**
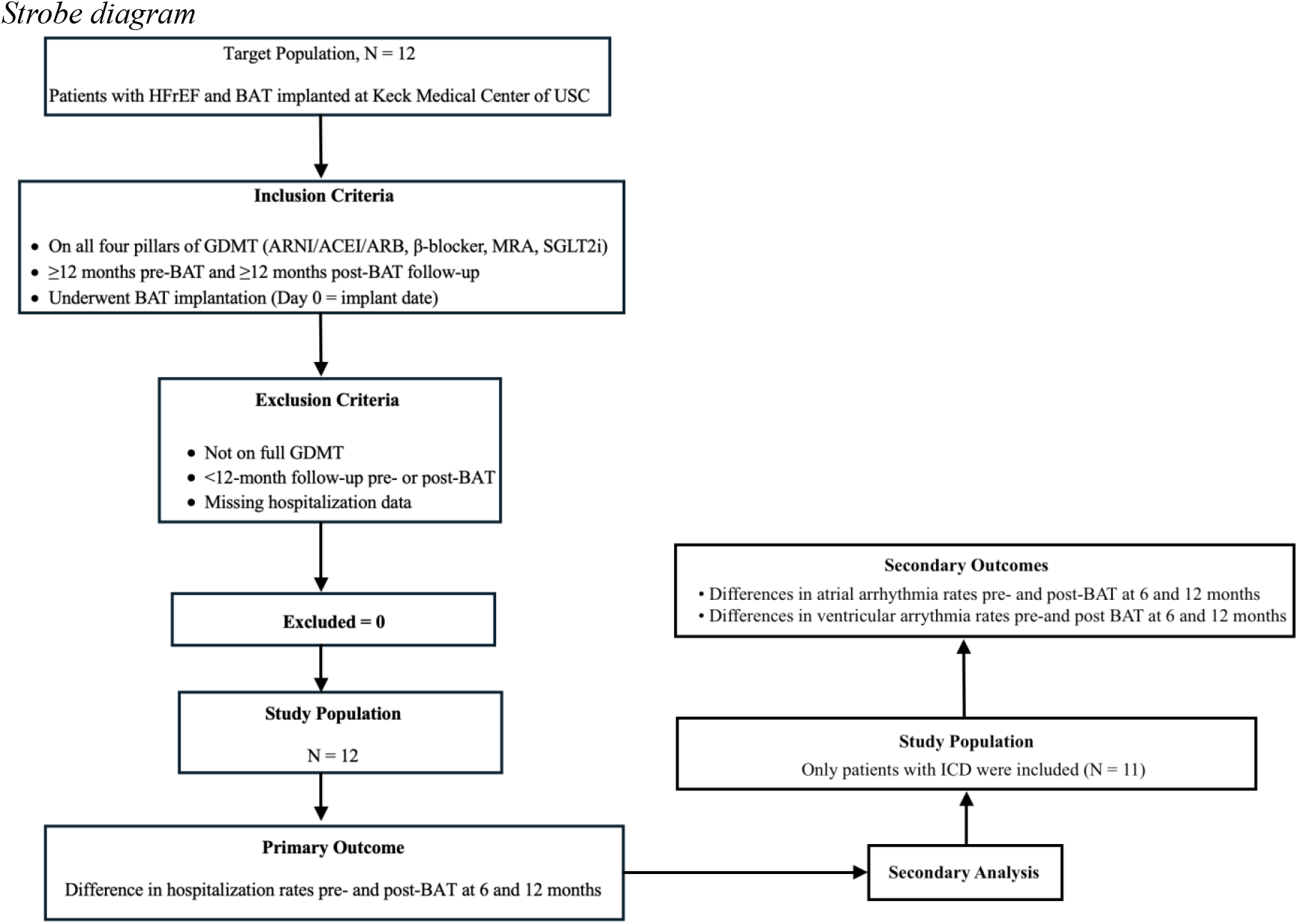
Strobe diagram

### Study Design

We analyzed hospitalization rates, AA rates, and VA rates for each subject 6 months and 12 months pre-BAT and post-BAT. Each patient served as their own control. The primary outcome was all-cause hospitalizations. For the secondary outcomes, patients with an ICD placed at least 12-months pre- and post-BAT therapy had their ICD interrogated every 1-3 months in clinic and each episode of AA and VA were recorded. VAs are defined as episodes of monomorphic and polymorphic VT, VF, and PVCs. AAs are defined as episodes of AF, AFL, and AT. Patients diagnosed with AA prior to the study timeline were excluded in the study. The data for AA and VA episodes are shown on Figures 2-5.

**Figure 2.**
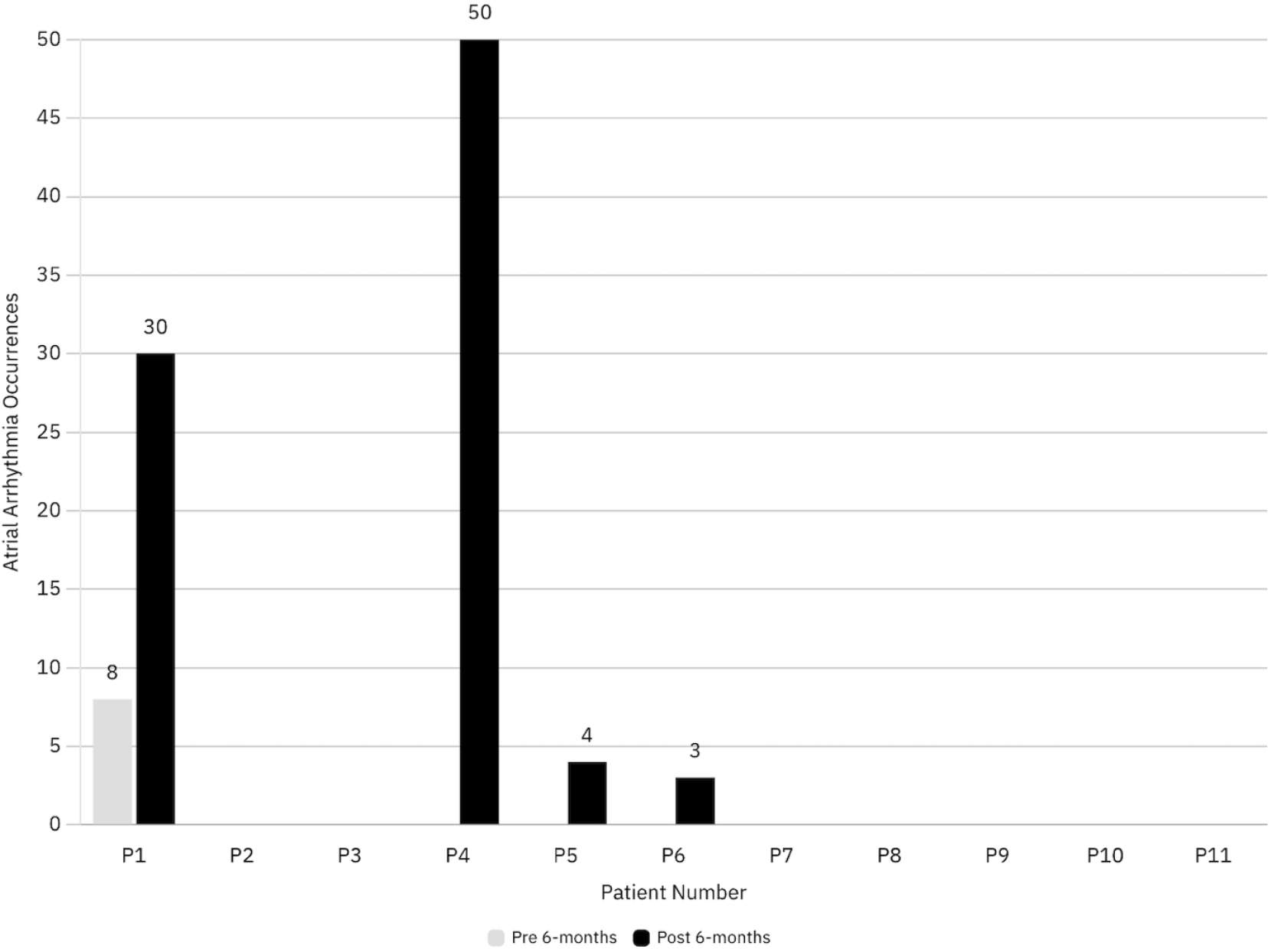
Atrial arrhythmia occurrences for each of the 11 patients 6-months pre- and post-BAT therapy.

**Figure 3.**
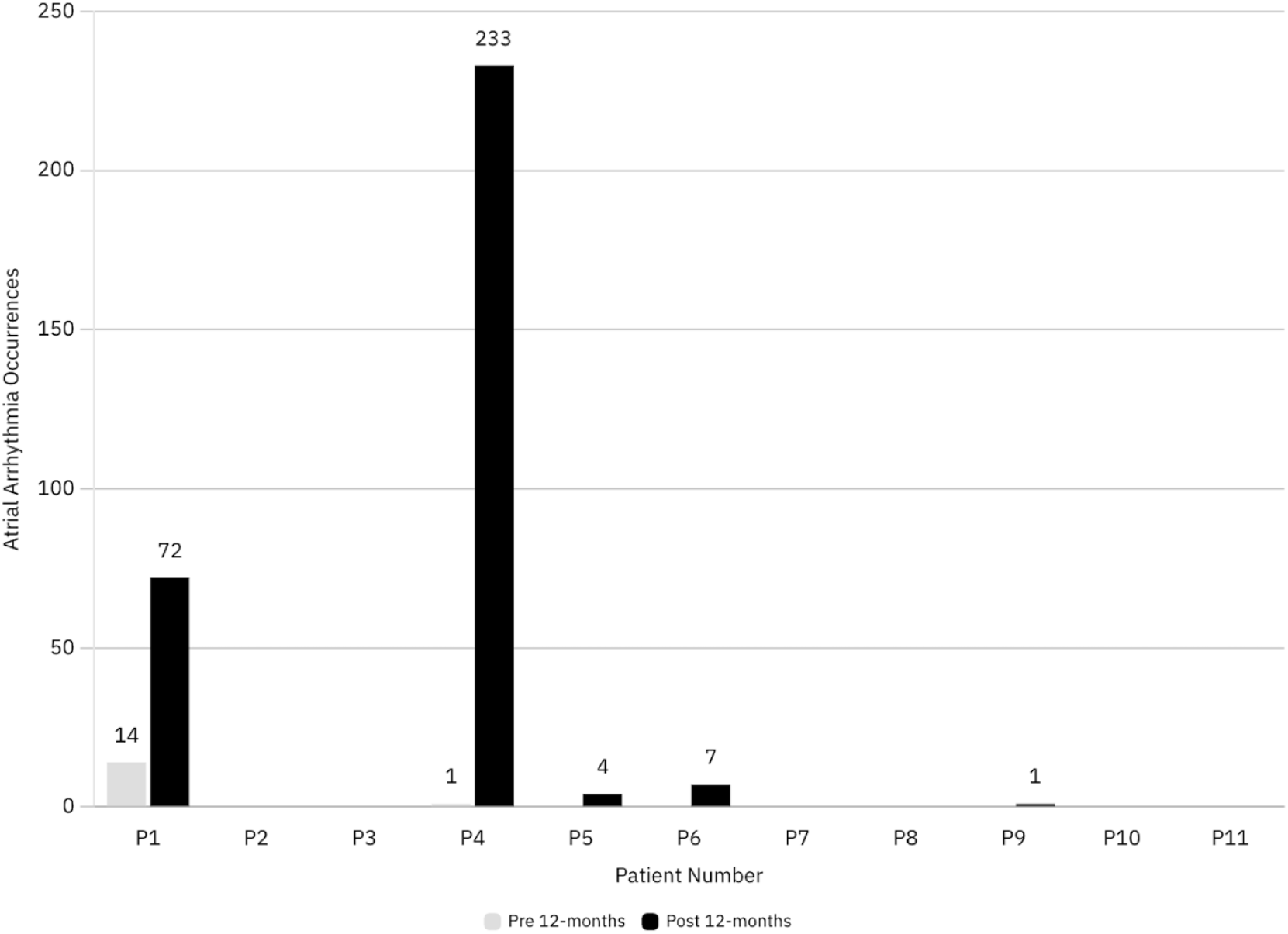
Atrial arrhythmia occurrences for each of the 11 patients 12-months pre- and post-BAT therapy.

**Figure 4.**
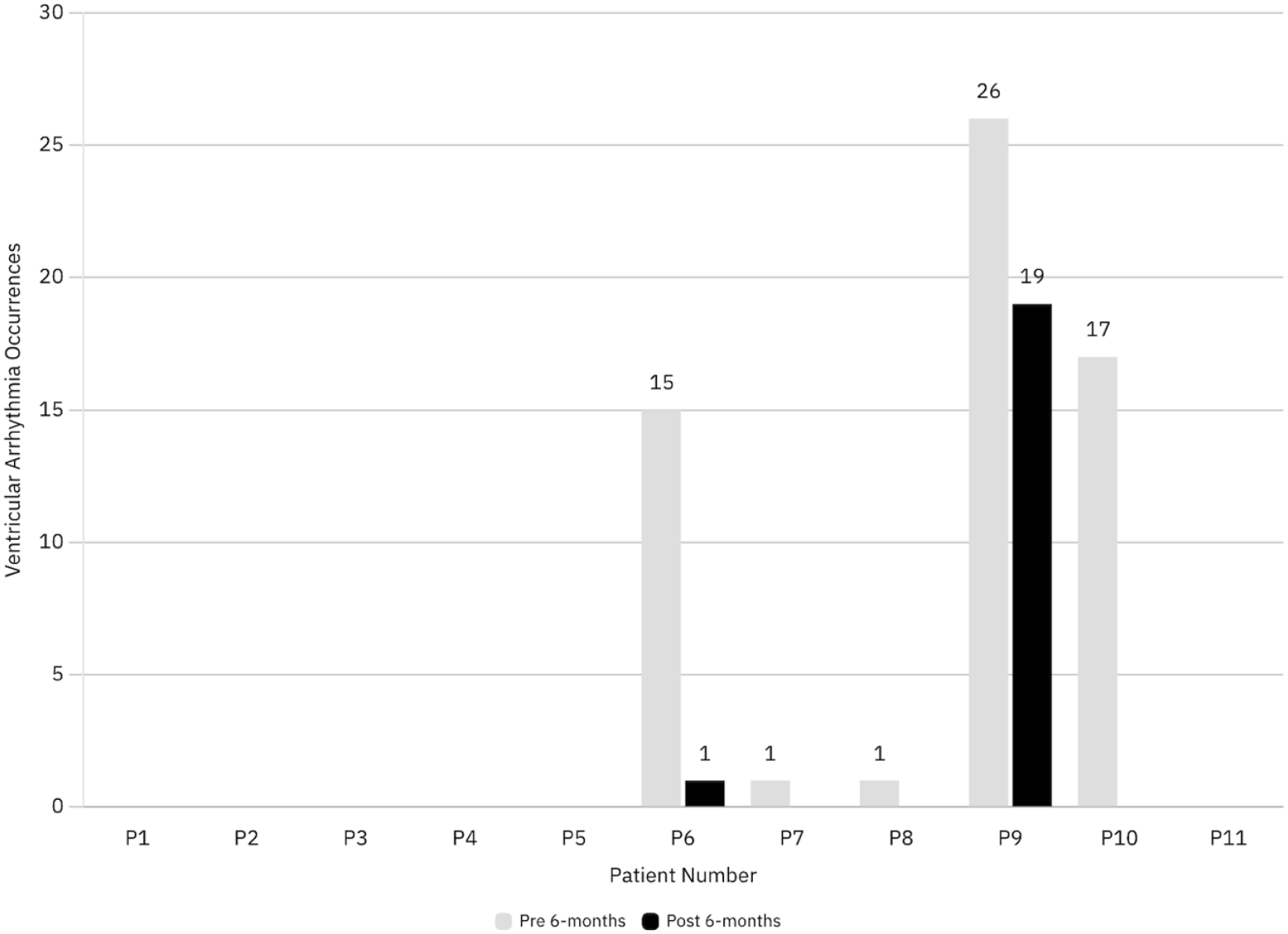
Ventricular arrhythmia occurrences for each of the 11 patients 6-months pre- and post-BAT therapy.

**Figure 5.**
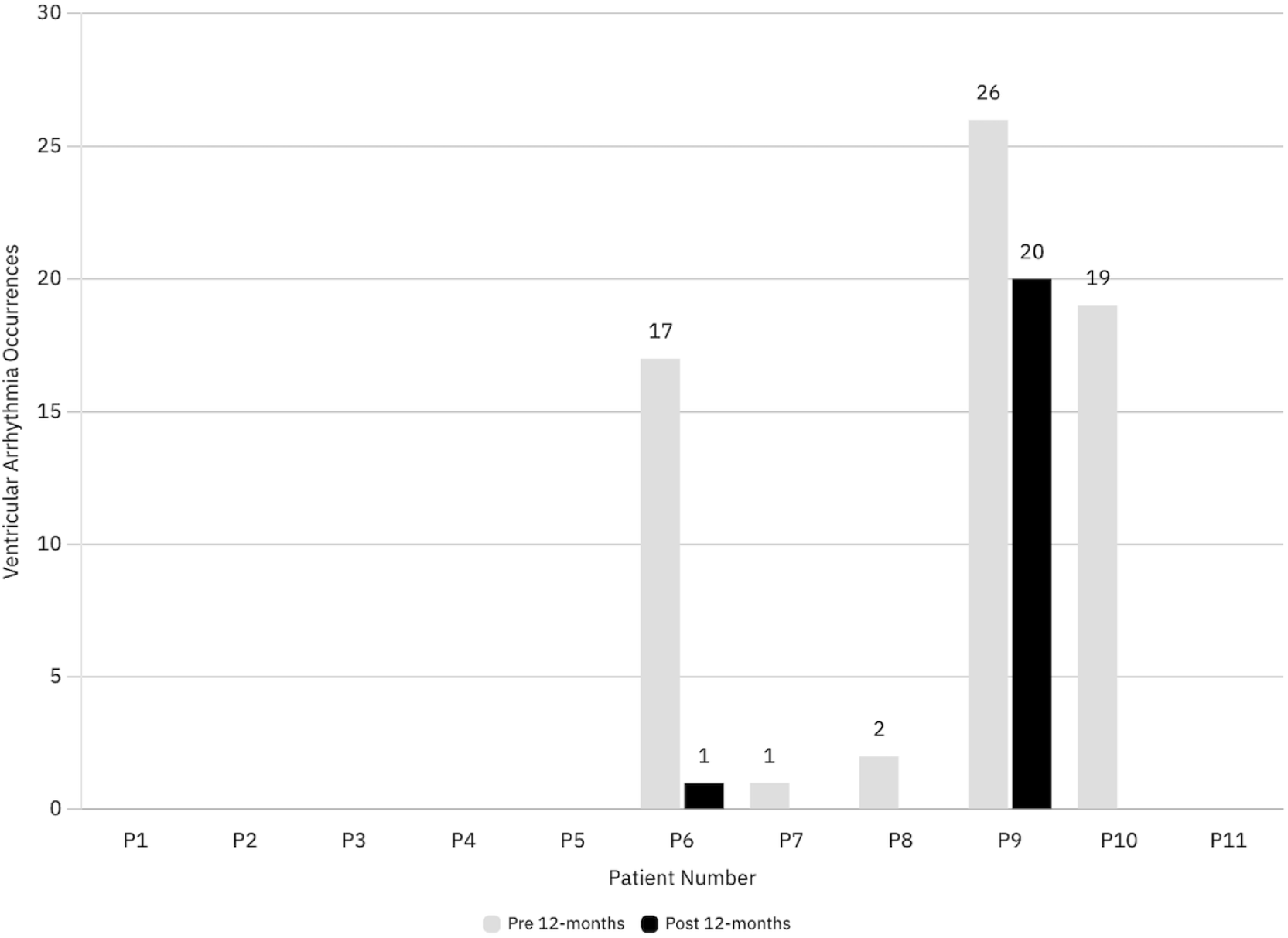
Ventricular arrhythmia occurrences for each of the 11 patients 12-months pre- and post-BAT therapy.

### Statistical Analysis

We used Wilcoxon Signed Rank tests to compare the primary outcome of hospitalization rates 6 and 12 months pre-BAT vs. post-BAT, and the secondary outcome of AA, and VA rates at the same time points. Specifically, 6-month pre-BAT rates were compared to 6-month post-BAT rates and 12-month pre-BAT rates were compared to 12-month post-BAT rates. As such, each patient served as their own control for both primary and secondary objectives. For ease of interpretation and comparison to existing literature, we present both means with standard deviation and median with interquartile range (IQR). A p-value ≤0.05 was considered statistically significant and no adjustments were made for multiple comparisons due to the exploratory nature of this study. Stata 18 (Statacorp, College Station, TX) was used to complete the analysis.

## Results

### Baseline Characteristics

The baseline characteristics of the 12 patients who met inclusion criteria are shown on Table 1. Baseline demographic and clinical characteristics are presented as mean and standard deviation for numeric variables and count with frequency for categorical variables. 31 patients were identified to be on BAT at the time of the analysis, and 12 patients fit the primary inclusion criteria of receiving all four pillars of GDMT for at least 12-months prior to BAT. Of those 12 patients, 11 patients fit the secondary outcome inclusion criteria of having an ICD placed at least 12-months pre-BAT therapy.

**Table 1.** Baseline characteristics for the 12 patients in the study.

| **VARIABLES (n=12)** | **MEAN ± SD or n (%)** |
| --- | --- |
| **AGE (years)** | 63.75 ± 14.75 |
| **SEX** |  |
| **MALES (n)** | 8 (67%) |
| **FEMALES (n)** | 4 (33%) |
| **DIABETES (n)** | 7 (58.3%) |
| **HYPERTENSION (n)** | 7 (58.3%) |
| **HYPERLIPIDEMIA (n)** | 10 (83.3%) |
| **CORONARY ARTERY DISEASE (n)** | 9 (75%) |
| **LEFT VENTRICULAR EJECTION FRACTION (%)** | 25.25 ± 10.79 |
| **HISTORY OF CIGARRETE USE (n)** | 5 (41.7%) |
| **CKD or ESRD (n)** | 5 (41.7%) |
| **CANDIDATE RACE (n)** | - |
| **HISPANIC** | 5 (41.7%) |
| **WHITE** | 7 (58.3%) |

### Hospitalization rates

The outcome comparison of hospitalization rates is shown on Table 2. There were no statistically significant differences between median pre- and post-BAT hospitalizations at 6 months [M_pre_ = 0 (IQR: 0, 1) vs. M_post_ = 0 (IQR: 0, 0), p = 0.63] (X_pre_ = 0.5 ± 0.9 vs X_post_ = 0.3 ± 0.9), however median hospitalizations were significantly higher pre-BAT than post-BAT at 12 months [M_pre_ = 1 (IQR: 0, 2.5) vs. M_post_ = 0 (IQR: 0, 0), p = 0.05] [X_pre_ = 1.3 ± 1.4 vs X_post_ = 0.3 ± 0.9.

**Table 2.** Comparison of Outcomes Between 6 and 12 months.

|  | Pre-BAT |  |  | Post-BAT |  |  |  |
| --- | --- | --- | --- | --- | --- | --- | --- |
| Time | n | Median (IQR) | Mean ± SD | n | Median (IQR) | Mean ± SD | p-value<sup>1</sup> |
| *Hospitalizations* |  |  |  |  |  |  |  |
| 6 Months | 12 | 0 (0, 1) | 0.5 ± 0.9 | 12 | 0 (0, 0) | 0.3 ± 0.9 | 0.63 |
| 12 Months | 12 | 1 (0, 2.5) | 1.3 ± 1.4 | 12 | 0 (0, 0) | 0.3 ± 0.9 | 0.05 |
| *Ventricular Arrhythmia* |  |  |  |  |  |  |  |
| 6 Months | 11 | 0 (0, 15) | 5.5 ± 9.3 | 11 | 0 (0, 0) | 1.8 ± 5.7 | 0.06 |
| 12 Months | 11 | 0 (0, 17) | 5.9 ± 9.7 | 11 | 0 (0, 0) | 1.9 ± 6.0 | 0.06 |
| *Atrial Arrhythmia* |  |  |  |  |  |  |  |
| 6 Months | 11 | 0 (0, 0) | 0.7 ± 0.4 | 11 | 0 (0, 4) | 7.9 ± 16.5 | 0.13 |
| 12 Months | 11 | 0 (0, 0) | 1.4 ± 4.2 | 11 | 0 (0, 7) | 28.8 ± 71.0 | 0.06 |

<sup>1</sup>P-values were obtained Wilcoxon Signed Rank tests.

### Atrial arrhythmia rates

The outcome comparison of AA rates is shown on Table 2. Median AAs did not differ pre- and post-BAT at 6 months [M_pre_ = 0 (IQR: 0, 0) vs. M_post_ = 0 (IQR: 0, 4), p = 0.13] (X_pre_ = 0.7 ± 0.4 vs X_post_ = 7.9 ± 16.5). In contrast, median AAs were borderline significantly different pre- and post-BAT at 12 months [M_pre_ = 0 (IQR: 0, 0) vs. M_post_ = 0 (IQR: 0, 7), p = 0.06] (X_pre_ = 1.4 ± 4.2 vs X_post_ = 28.8 ± 71.0).

### Ventricular arrhythmia rates

The outcome comparison of VA rates is shown on Table 2. There were more VA at both 6 months pre and post-BAT [M_pre_ = 0 (IQR: 0, 15) vs. M_post_ = 0 (IQR: 0, 0)], p = 0.06] (X_pre_ = 5.5 ± 9.3 vs. X_post_ = 1.8 ± 5.7) and 12 months pre and post-BAT [M_pre_ = 0 (IQR: 0, 17) vs. M_post_ = 0 (IQR: 0, 0), p=0.06] (Xpre = 95.9 ± 9.7 vs. Xpost = 1.9 ± 6.0) respectively), however, these differences failed to reach statistical significance.

## Discussion

In patients with HFrEF on the current-era GDMT, our findings suggest that median hospitalization rates are lower with BAT at 12-months. We found a borderline statistically significant increase in AA rates at 12-months, and borderline statically significant reduction in VA rates at 6-months and 12-months. Combined with other available literature, our analysis has shown that BAT may be beneficial in patients with HFrEF in conjunction with maximally tolerated GDMT.

Previous analyses have shown that BAT improves HF symptoms through improvements in 6-min walk distance, quality-of-life score, and NYHA functional class ranking.^3,10,11^ These studies compared patients receiving GDMT alone with those receiving GDMT plus BAT.^3,10,11^ In contrast, our analysis used a within-subject design, treating each patient as their own control to minimize confounding factors. Furthermore, due to the timing of the prior studies, the most recent GDMT additions of ARNIs and SGLT-2 inhibitors may not have been available.^3,10,11^ Our study exclusively evaluated patients already on current era GDMT, which includes all four pillars of medication classes shown to improve mortality in patients with HFrEF, prior to BAT.^12^

BAT is effective in treating drug-resistant hypertension with a prior study showing decreased frequency and duration of hypertension-related hospitalizations.^13,14^ A more recent study reported shorter hospital stays and reductions in all-cause, cardiovascular, and HF-related hospitalizations associated with BAT.^15^ Our study, which focused on all-cause hospitalizations, is consistent with these findings which further supports the beneficial impact of BAT in patients with HFrEF.

Finally, our study analyzed the effects of BAT on atrial and ventricular arrhythmia rates. Patients with HFrEF are at high risk of developing AAs and VAs, which may ultimately lead to increased morbidity and mortality.^5-7, 17^ While a reduction an atrial and ventricular arrhythmias has been reported by others, our study showed a borderline significant increased AA rates at 12-months post-BAT and borderline significant reduced VA rates at 6 and 12-months post-BAT,^16^ though these findings did not reach statistical significance.

### Limitations

These findings should be considered in the context of several limitations. Foremost, our sample size is small, owing to the single-center design and rarity of BAT use in HFrEF. As a result, we may have missed some true differences. The small sample size also contributes to increased variability in the measurements, leading to a lower likelihood of a significant result. Also resulting from the limited sample, we were unable to evaluate and adjust for possible confounding variables, though the matched design reduces the risk of confounding. Last, we note that these results may not generalize outside of the study population and thus, additional studies are needed to provide more clarity. In addition to increasing sample size, future studies in this area should address whether benefits persist many years for patients with HFrEF.

## Conclusions

In conclusion, we observed reduced hospitalization rates at 12-months post-BAT therapy compared to 12-months pre-BAT therapy in HF patients on all four classes of GDMT, with borderline significant increases in AA rates at 12 months and decreasing VA rates at 6 and 12 months of BAT therapy.

## Data Availability

All data obtained in the manuscript are available upon request.

## Abbreviations

GDMT: Guideline-Directed Medical Therapy
HF: Heart Failure
HFrEF: Heart Failure with Reduced Ejection Fraction
HFmrEF: Heart Failure with Mid-Range Ejection Fraction
BAT: Baroreflex Activation Therapy
ICD: Implantable Cardioverter-Defibrillator
VA: Ventricular Arrhythmias
VT: Ventricular Tachycardia
VF: Ventricular Fibrillation
PVC: Premature Ventricular Contractions
EMR: Electronic Medical Record

